# Water, Sanitation and Hygiene Interventions in Schools for Effective Pandemic Response in Low and Middle-Income Countries: Scoping review protocol

**DOI:** 10.1101/2023.08.09.23293884

**Authors:** Gladys Chepkorir Seroney, Gugu Gladness Mchunu, Kabelo Kgarosi, Ng’wena Asha Gideon Magak

## Abstract

**Background:** The impact of improved water, sanitation, and hygiene (WASH) access on mitigating illness is well documented. Despite continued national and international efforts, access to improved water and sanitation in schools remains limited in many developing countries. The proposed scoping review is aimed at mapping evidence on the status of WASH interventions in schools for effective pandemic response in low and middle-income countries. The scoping review will guide and improve schools’ WASH with an ultimate goal of preventing disease and protecting school-going children during infectious disease outbreaks, including the current COVID-19 pandemic.

**Methods:** The proposed scoping review will be guided by Arksey and O’Malley’s framework. A comprehensive keyword search for relevant articles presenting evidence on the status of WASH in school interventions will be conducted in the Cumulative Index to Nursing and Allied Health Literature (CINAHL), PubMed, Academic Search Complete, Google Scholar, and the Scopus electronic databases. Articles reporting on the status of WASH in schools published between January 2000 and September 2021 will be included. The review will employ the NVIVO version 12 software package to extract the relevant themes from the included articles using content thematic analysis.

**Discussion:** We anticipate to find relevant studies reporting on the status of WASH interventions in schools. The results of this review will provide information that is likely to inform the monitoring and evaluation of WASH interventions in schools and assist towards achievement of Sustainable Development Goal 6. It may also help in crafting relevant and up to date guidelines or policies in relation to WASH systems in schools.

## INTRODUCTION

Water, Sanitation and Hygiene (WASH) are fundamental to promoting child health in low and middle-income countries. Water scarcity, limited access to improved sanitation and lack of personal hygiene at home and in school significantly contribute to the immense burden of preventable childhood diseases, such as diarrhoea, acute respiratory infections, intestinal worms and dental caries (1). According to Gwenzi (2) the need for clean water provision, sanitation and hygiene has only received limited attention in developing countries, while provision of clean drinking water, proper sanitation, food safety and hygiene could be critical in the current fight against COVID-19. WASH have been identified as key determinants of community and public health status in human populations. The United Nations International Children’s Emergency Fund (UNICEF) formulated the WASH interventions in schools which aimed at improving the health and hence academic performance of children in all regions of the world (3).

During infectious disease outbreaks, such as the current COVID-19 pandemic, well-managed WASH facilities are critical for preventing disease and protecting public health. Investing in core public health infrastructure, such as water supply and sanitation systems, is one of the most cost-effective strategies for pandemic preparedness, particularly in resource-constrained contexts (4, 5, 6) Schools’ WASH programs are being scaled-up globally while little is known about the actual status of resources in the low income regions and how such programs are being monitored and evaluated (7).

UNICEF and WHO have developed guidelines for minimum standards for WASH in schools. The United Nations General Assembly (UNGA) in its recognition of the human right to water and sanitation in the year 2010 supported the WASH policy (8). In addition, currently, the Sustainable Development Goal (SDG) 6 aims to ensure availability and sustainable management of water and sanitation for all. The goal comprises of six technical targets relating to drinking water, sanitation and hygiene, waste water management, water efficiency, integrated water resource management and protection of the aquatic ecosystems (9)

The protection of children and educational facilities is particularly important. Precautions are necessary to prevent the potential spread of COVID-19 and other infectious diseases in school settings (10, 11)

The proposed scoping review study seeks to map evidence on the status of water, sanitation and hygiene interventions in schools for effective pandemic response. Pandemics are communicable diseases which include COVID 19, Ebola, Influenza, Avian and swine flu, plague, cholera, Rift Valley fever and yellow fever, to mention but a few. It is anticipated that the outcome of the study will be a basis for improving school safety during the COVID-19 pandemic and beyond.

The findings of this study will provide information to governments on the current status of water, sanitation and hygiene interventions in schools. The result will also provide information to UNICEF and WHO that will assist in monitoring the progress of WASH in schools, and also in the achievement of Sustainable Development Goal 6 (11)

It will also help them prioritize on the most vulnerable schools that require school specific tailored interventions. It is anticipated that the findings of this proposed study will provide insights to the community on how best to protect and promote good health status of both the school-going children and the community at large.

## Methodology

A scoping review of water, sanitation and hygiene will be conducted mainly to assess the status of WASH interventions in schools and their levels of preparedness for effective pandemic response. The scoping review will be guided by Arksey and O’Malley’s (12) framework for conducting scoping reviews and mapping evidence. The framework includes the following stages: identification of the research question, identification of relevant studies, study selection, charting the data, and collating, summarizing, and reporting of the results (13).

A PRISMA-ScR checklist will be attached as part of supplementary material (Fig 1). The PRISMA-ScR checklist contains 20 essential reporting items and 2 optional items. The intent of the PRISMA-ScR is to help readers (including researchers, publishers, commissioners, policymakers, health care providers, guideline developers, and consumers) develop a greater understanding of relevant terminology, core concepts, and key items to report for scoping reviews. The Joanna Briggs Institute (JBI) endorsed Preferred Reporting Items for Systematic Reviews and Meta-Analyses for Scoping Reviews (PRISMA-ScR) checklist will be completed in order to structure the reporting of the scoping review (14).

**Figure 1.**
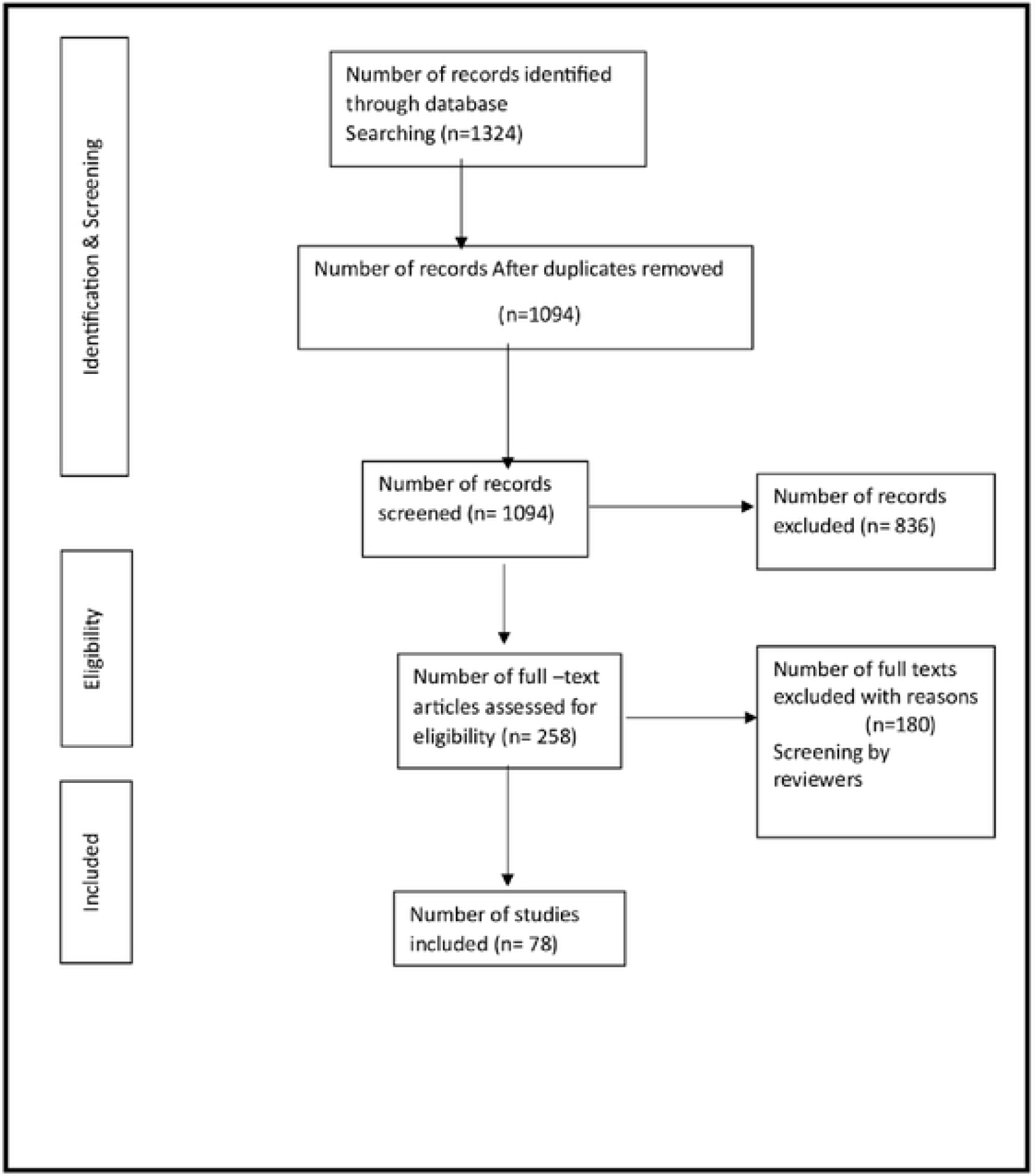
PRISMA extension for scoping reviews, 2018 flow diagram

The proposed scoping review is registered on OSF (Open Science Framework) and can be accessed via this link: https://osf.io/rytp9

### Stage 1: Identify the research questions

The scoping review will seek to answer the research question: W*hat is the status of Water, Sanitation and Hygiene interventions in schools for effective response to disease pandemic?*

To guide the research question, the search will utilize the JBI PCC mnemonic: Population, Concept and Context (15) presented in table 1.

**Table 1:** The PCC mnemonic.

| PCC mnemonic | Inclusion criteria |
| --- | --- |
| Population | Primary Schools |
| Concept | Water, Sanitation and Hygiene |
| Context | Diseases Pandemics/ infectious diseases |

### Stage 2: Identifying relevant studies

A comprehensive search of relevant articles from the following electronic databases, including PubMed, Cumulative Index to Nursing and Allied Health Literature (CINAHL), Academic Search Complete, Google Scholar, and Scopus will be conducted for articles published from January 2000 to August 2021.

A summary of the key word search strategy used to answer our research question using the scoping research methods is outlined in table 2.

**Table 2:**
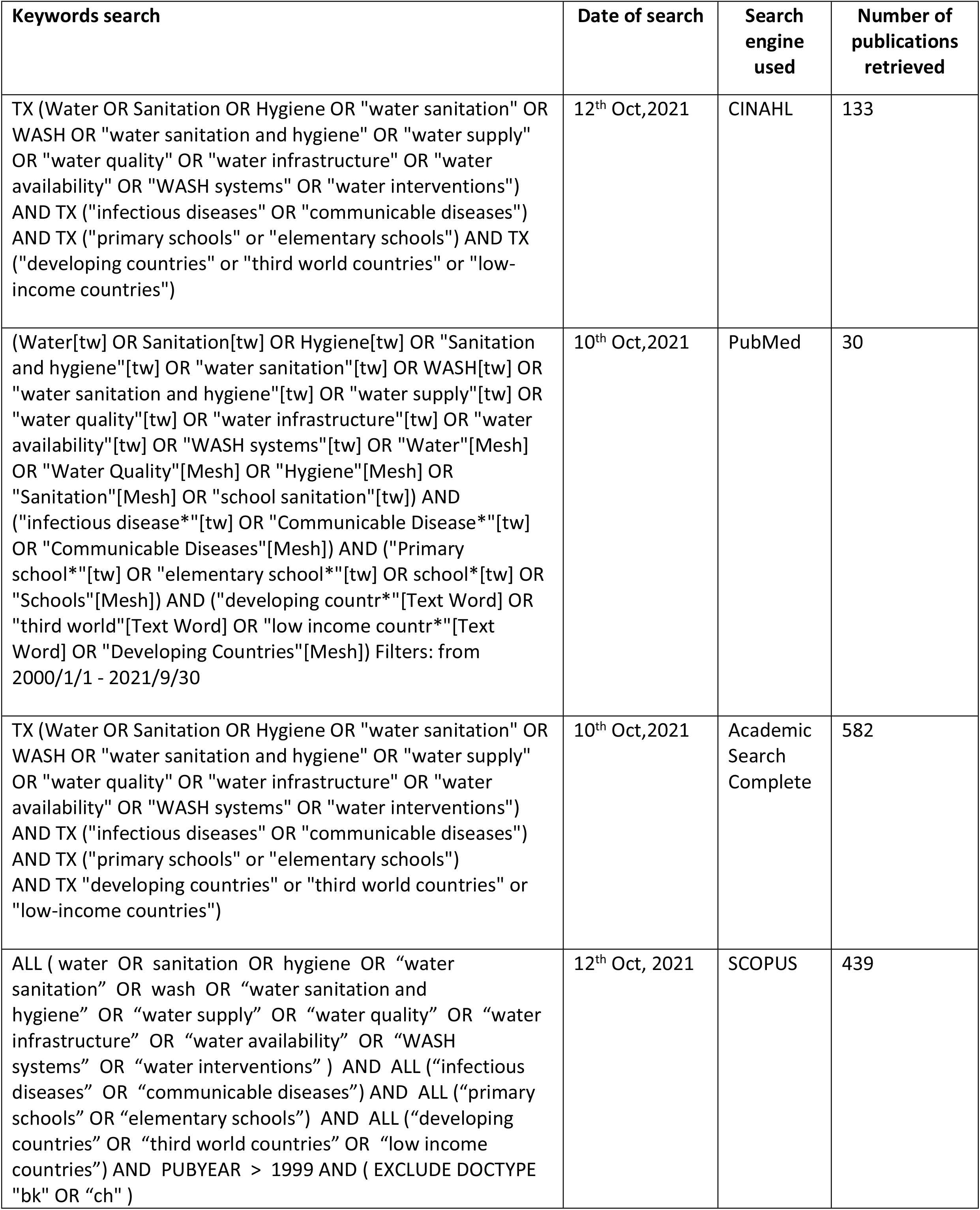
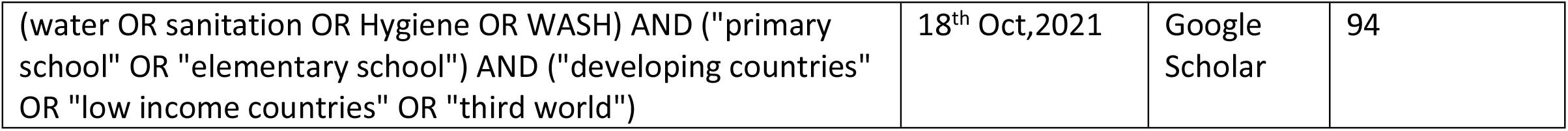
Key words search strategy and databases.

### Stage 3: Study Selection

The following eligibility criteria were developed to ensure inclusion of relevant evidence to help us answer the scoping review research question.

We will apply the following eligibility criteria to ensure a rigorous evidence review process.

#### Inclusion criteria

- Articles that focus solely on WASH in schools
- Studies that address WASH in schools and communicable diseases/ disease outbreaks/ disease pandemics
- All studies/articles published in English and other languages that are relevant to the study
- Peer-reviewed literature
- No methodological restriction
- Articles/ studies published from January 2000 to September 2021

#### Exclusion criteria

- Articles that focus on communicable diseases/ disease outbreaks/ disease pandemics
- without reference to WASH in schools
- Articles addressing WASH outside schools

The review process will be conducted in three stages of screening. The first screening stage is where the Principal Investigator (PI) will screen the titles of the articles retrieved from the database searches. An Endnote library using Endnote20 will be created, and all eligible articles following title screening will be exported into the library. The second screening will be conducted by two independent trained reviewers who will screen abstracts in parallel. Any disagreement in screening between the two reviewers will be resolved by discussion and consensus. The third screening will involve screening of full text articles that became eligible following the abstract screening; this screening will also be done by two independent trained reviewers (12). Kappa statistics will be calculated to determine the level of agreement between the two reviewers. The kappa statistic of > 0.61 will be considered acceptable agreement (16). Discrepancies in reviewers’ responses following full article screening will be resolved by inviting a third screener who should be the study PI or the team’s senior research member. We will report the screening results following the Preferred Reporting Items for Systematic Reviews and Meta-Analyses (PRISMA) guidelines (Fig 2).

### Stage 4: Charting the data

A data-charting form will be developed and used to extract data from each article in order to determine whether the independent researchers are utilizing an approach that is consistent with the research question. Two independent trained reviewers will pilot the data charting form using a random sample of some included studies for consistency. Modification of the data extraction form will be done as required based on the feedback from the two independent reviewers. The data charting form will be constantly updated throughout the duration of the study. The variables to be included in the checklist are the author, year of publication, study title, the aim of the study, study population, study setting, study results or findings, and conclusions (13). (Table 3)

**Table 3:**
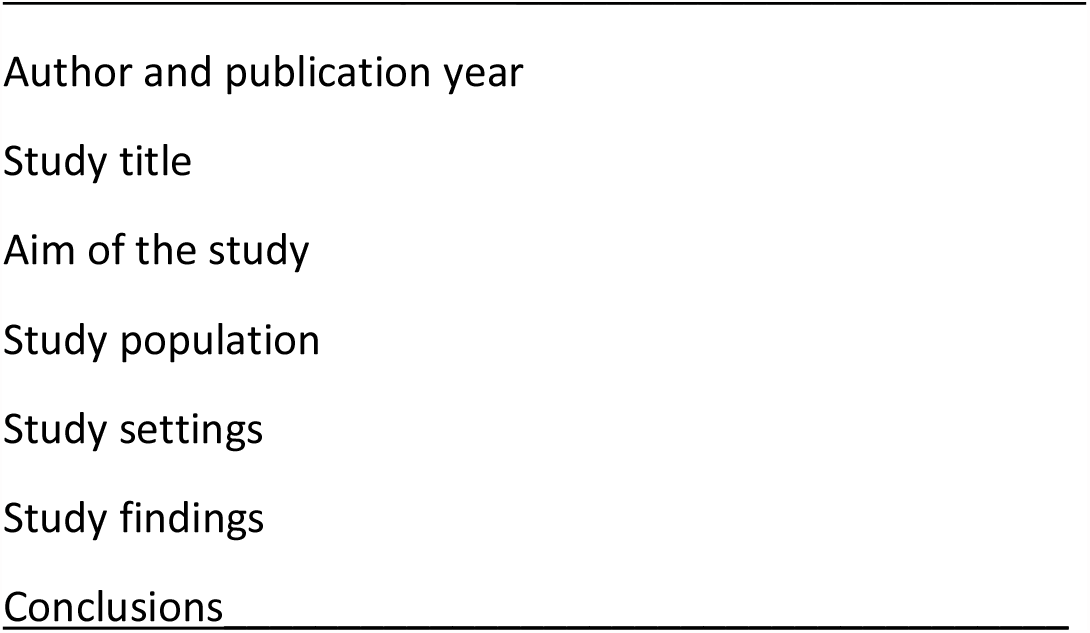
Data charting form.

Rayyan, a free web-tool (Beta) designed to help researchers working on systematic reviews and scoping reviews, will be used to screen and select studies after importing them from EndNote

### Stage 5: Collating, summarizing and reporting the results

NVivo version 12 will be employed to conduct content thematic analysis of the included studies. The thematic content analysis steps include the following: familiarizing with data, generating initial codes, searching for themes, reviewing themes, defining and naming themes, and producing the report (17). The findings presenting the main concepts from the included articles in line with the research question, will be presented in a narrative format then published in a peer reviewed journal.

#### Quality appraisal

We will conduct quality appraisal of the included articles using the mixed methods appraisal tool (MMAT) 2018 version. The tool will guide us through 4 steps; firstly, apply the screening questions for all studies; secondly, for each relevant study determine the type of design, and use the corresponding criteria to appraise the study’s quality; thirdly, two independent reviewers conduct the appraisal process; and, lastly, determine an overall quality score for each study.

#### Ethical considerations

This study will not include human or animal participants; therefore, it does not require ethical approval.

#### Patient and Public Involvement

The study will not involve patient and public involvement since it is a scoping review where the researcher will review previous literature from credible sources to identify available evidence of WASH interventions in school.

## Discussion

The proposed scoping review is aimed at mapping evidence on WASH systems in schools in order to guide and improve schools’ WASH with a goal of preventing disease and protecting school-going children during infectious disease outbreaks, including the current COVID-19 pandemic. Addressing WASH is important because the global community has committed to achieving 17 Sustainable Development Goals (SDGs) by 2030, of which Goal 6 which states that access to safely managed WASH should be universally available and is a global priority (18).

Articles that focus on infectious diseases without refence to WASH in schools will be excluded from the review because the review is to map literature and show evidence of the status of WASH interventions in schools specifically, not WASH in general. Articles on infectious diseases that do not have reference to WASH in schools will not be included as they do not make any objective contribution towards the aim of the review.

It is anticipated that the result of this review will provide information to UNICEF and WHO in monitoring the progress of WASH in schools, and also in the achievement of Sustainable Development Goal 6 (11). The findings may also help the Ministry of Health in crafting relevant and up to date guidelines or policies in relation to WASH interventions in schools. The results will also give insight on further areas of research concerning WASH in schools in order to protect school-going children from pandemics.

## Data Availability

The data of this study will be made fully available and without restriction at the repository of Durban University of Technology Library. South Africa. Or anywhere recommended by the PLOS Global Public Health

## Supplementary information

CINAHL- Cumulative Index to Nursing and Allied Health Literature

JBI- Joanna Briggs Institute

MMAT- Mixed Methods Appraisal Tool (MMAT)

PRISMA- ScR- Preferred Reporting Items for Systematic Reviews and Meta-Analyses for Scoping Reviews

SDGs- Sustainable Development Goals

UNGA- United Nations General Assembly (UNGA)

UNICEF- United Nations Children’s Fund

WASH- Water, Sanitation and Hygiene

WHO- World Health Organization

## Declarations

## Acknowledgements

The authors would like to extend their appreciation to UNICEF and the Faculty of Health Sciences, University of Pretoria, for Evidence Synthesis Capacity Building for Women Scientists in Sub-Saharan Africa that has supported the development of this research study.

Durban University of Technology and University of Pretoria library for availing resources.

## Author’s information

Gladys Chepkorir Seroney is the Head of Community Health Nursing Department in the School of Nursing, Maseno University, Kisumu, Kenya.

## Ethical approval and consent to participate

Not applicable

## Consent for publication

Not applicable

## Availability of data and materials

All data generated or analyzed during this study will be included in the published scoping review article

## Competing interests

The authors declare that they have no competing interests.

## Authors’ contributions

GCS conceptualized the study and prepared the draft proposal under the supervision of GGM and MADN. GCS contributed to the development of the background as well as designing the study and manuscript preparation. GSC and KK participated in accessing library resources and search strategy. All authors (GCS, GGM, MADN, and KK) contributed to the review, editing of the manuscript and approved the final version.

## Funding

The training on Evidence synthesis has been made possible by the funding from UNICEF and with support from Future Africa, the University of Pretoria.

## Notes

### Competing Interest Statement

The authors have declared no competing interest.

### Funding Statement

The training on Evidence synthesis on how to conduct systematic reviews has been made possible by the funding from UNICEF and with support from Future Africa, the University of Pretoria.

### Author Declarations

This is a scoping review protocol. It does not require participants consent

